# Activated NK Cells with Pro-inflammatory Features are Associated with Atherogenesis in Perinatally HIV-Acquired Adolescents

**DOI:** 10.1101/2023.11.06.23297580

**Authors:** Mario Alles, Manuja Gunasena, Aaren Kettelhut, Kate Ailstock, Victor Musiime, Cissy Kityo, Brian Richardson, Will Mulhern, Banumathi Tamilselvan, Michael Rubsamen, Dhanuja Kasturiratna, Thorsten Demberg, Cheryl M. Cameron, Mark J. Cameron, Sahera Dirajlal-Fargo, Nicholas T. Funderburg, Namal P.M. Liyanage

## Abstract

Human immunodeficiency virus (HIV) is associated with persistent immune activation and dysfunction in people with HIV despite treatment with antiretroviral therapy (ART). Modulation of the immune system may be driven by: low-level HIV replication, co-pathogens, gut dysbiosis /translocation, altered lipid profiles, and ART toxicities. In addition, perinatally acquired HIV (PHIV) and lifelong ART may alter the development and function of the immune system. Our preliminary data and published literature suggest reprogramming innate immune cells may accelerate aging and increase the risk for future end-organ complications, including cardiovascular disease (CVD). The exact mechanisms, however, are currently unknown. Natural killer (NK) cells are a highly heterogeneous cell population with divergent functions. They play a critical role in HIV transmission and disease progression in adults. Recent studies suggest the important role of NK cells in CVDs; however, little is known about NK cells and their role in HIV-associated cardiovascular risk in PHIV adolescents. Here, we investigated NK cell subsets and their potential role in atherogenesis in PHIV adolescents compared to HIV-negative adolescents in Uganda. Our data suggest, for the first time, that activated NK subsets in PHIV adolescents may contribute to atherogenesis by promoting plasma oxidized low-density lipoprotein (Ox-LDL) uptake by vascular macrophages.

## Introduction

HIV remains a significant public health challenge, particularly in pediatric and adolescent populations. According to the Joint United Nations Program on HIV/AIDS (UNAIDS), there were an estimated 1.5 million children (aged 0-14 years) and 1.1 million adolescents (aged 15-19 years) living with HIV globally in 2022 (UNAIDS, 2023 estimates). The majority (almost 86%) of these children reside in the Sub-Saharan African region where the primary mode of transmission for those under 15 years of age is perinatal transmission (1). With access to ART, however, these children are able to live well into adulthood (2). HIV and its treatment with ART are associated with persistent immune activation and immune dysfunction (3). This may be driven by low level HIV replication, co-pathogens, gut microbial translocation and ART toxicity (4). This persistent immune dysregulation may be a culprit in the increased incidence of non-AIDS related co-morbidities including cardiovascular diseases, neurocognitive disorders, and cancer (5–9). CVD is the most common cause of morbidity and mortality in people living with HIV (PLWH) (6, 10–13) and has been found to be up to three times more common in these individuals when compared to those without HIV (14, 15). Specifically, PLWH have higher rates of coronary artery disease and atherosclerosis, an asymmetrical focal thickening of the intima of arteries with lipid plaque, which can result in myocardial infarction (16). So far, the primary immune cells linked to the inflammatory process surrounding atherosclerotic plaque development, or atherogenesis, are monocytes/macrophages (17). Other immune cells, including T lymphocytes likely contribute to this phenomenon, and the characterization of immune cell contributions to atherogenesis could have significant therapeutic implications (18–20).

NK cells are a group of innate lymphoid cells that are important in the surveillance and clearance of viral infections and play a key role in the immunopathogenesis of HIV (21, 22). NK cells are critical mediators of the innate immune response and are especially important in the pediatric population, where they take precedence during pathogen challenge over a still-developing adaptive immune response in early childhood (23). Even though traditionally thought to be immune cells with purely innate immune features, there is now mounting evidence that human NK cells display features of antigen specificity and memory, as depicted by their vaccination-dependent antigen-specific recall responses (24). Such “recall responses” may be related to a recent concept known as trained immunity, where innate immune cells are epigenetically reprogrammed to respond differently on subsequent exposure to a microbial product. Trained immunity can be induced by microbial products such as lipopolysaccharide and 1,3-β-D-glucan, which frequently enter the blood stream because of increased gut permeability in PLWH (25).

NK cells can be classified into different subtypes based on the relative expression of CD56 and CD16 receptors and by their different functional capabilities, with higher expression of CD56 signifying increased cytokine producing potential (CD56^bri^CD16^dim/-^), and higher expression of CD16 (CD56^dim/-^CD16^bri^) reflecting more cytotoxic capabilities (26). Among these subpopulations, varying degrees of expression of activating (e.g., CD69, CD57, NKp44, HLA-DR) and inhibitory (e.g., NKG2A) receptors and cytokine/chemokine receptors (e.g., CCR5, CXCR3, CXCR5) determine their functional and migratory characteristics (27).

Studies have demonstrated that HIV differentially affects protein expression among several NK cell subsets (26). A study among a cohort of neonates in Botswana with perinatally acquired HIV revealed that CD57^+^CD56^dim^CD16^dim^ NK cells, cells with elevated antiviral and cytotoxic properties, increased over time following birth (28). This increase correlated negatively with the frequency of intact proviruses, signifying that this subpopulation of cells played a role in reducing HIV-1 reservoir cells (29). Furthermore, among PLWH, these activated cytotoxic cells can express chemokine receptors like CCR5 and CXCR4 through trogocytosis, thereby enhancing their pro-inflammatory potential (30).

While substantial evidence exists regarding the significance of NK cell subsets in HIV among adults and children, a research gap exists in understanding their role in adolescents with PHIV exposed to HIV and/or ART since birth. Emerging concerns suggest that HIV’s impact on adolescents may surpass that in adults due to heightened inflammation and prolonged immune activation. This sustained immune dysregulation in PHIVs may be dominated by NK cells with altered phenotypic and functional characteristics. Given the role of NK cells in atherosclerosis (31), we hypothesize that persistent NK cell activation/alteration in PHIV may contribute to atherogenesis in PHIVs. In this study, we found that PHIV adolescents exhibit activated and pro-inflammatory NK cell profiles associated with HIV, which directly influence Ox-LDL uptake by macrophages, driving foam cell (lipid-laden macrophages) formation and atherogenesis. Additionally, we identify NKG2D/ligand interaction as a potential mediator of this mechanism. These findings highlight novel therapeutic and preventive targets to mitigate cardiovascular disease risk in this patient group.

## Result

### Study design and cohort demographics

For our analysis, we used cryopreserved peripheral blood mononuclear cells (PBMC) and plasma samples obtained from two groups: adolescents living with perinatally acquired HIV (PHIV) (n=18) and a group of age– and sex-matched adolescents unexposed and HIV negative (HIV-, n=20). These specimens were collected during a study conducted at the Joint Clinical Research Center (JCRC) in Kampala, Uganda, between 2017 and 2021 as previously described(32, 33). Both PHIV and HIV-adolescents were prospectively enrolled in this observational cohort study. Overall, the mean age was 14 years (SD=1.7), 52 % were females, and 94% PHIV participants had undetectable viral load (<50 copies/mL). Amongst PHIVs, about 53% were on a dolutegravir-based regimen, the remaining were on nevirapine, efavirenz or the boosted protease inhibitor lopinavir/ritonavir due to the ongoing transition to the newer WHO ART guidelines at the time of the participant enrollment (34). We did not observe any significant differences in CMV seropositivity between the two groups (Table 1). No significant differences were also observed in the routinely measured serum lipids, including cholesterol, high-density lipoprotein, low-density lipoprotein, very low-density lipoprotein, and triglycerides between PHIVs and HIV-(Table 2).

### NK cells in HIV-unexposed adolescents show high receptor diversity

To determine the impact of HIV on NK cell subsets in PHIV, we first examined cell populations among HIV-unexposed adolescents using high-dimensional spectral flow cytometry in conjunction with conventional flow cytometry analysis (Supplemental Figure 1A). Additionally, we utilized FlowSOM-based automatic clustering algorithms, and Uniform Manifold Approximation and Projection (UMAP) for dimensional reduction visualization (Figure 1A). This analysis was conducted on CD45^+^ CD3^−^ CD19^−^ cells to discern NK cell clusters and subsets within the cohort. We next conducted a clustered heatmap analysis (Figure 1B), revealing the presence of eight distinct NK cell clusters and two monocyte cell clusters. Furthermore, when applying the previously described gating strategy based on CD56 and CD16 expression to identify NK cells in neonates (28), we identified six distinct NK cell subsets in HIV-adolescents (Figure 1C); the prominent NK cell population in the blood of adolescents without HIV was CD56^dim^CD16^dim^, followed by CD56^−^CD16^dim^, and CD56^dim^CD16^−^ (Figure 1D). CD16^+^ NK subsets displayed a more activated phenotype, with increased expression of activation markers such as HLA-DR (Figure 1H) (35). In contrast, the CD56^+^ NK subsets showed a more inhibitory phenotype, marked by elevated expression of the inhibitory marker NKG2A (Figure 1K) (36).

**Figure 1.**
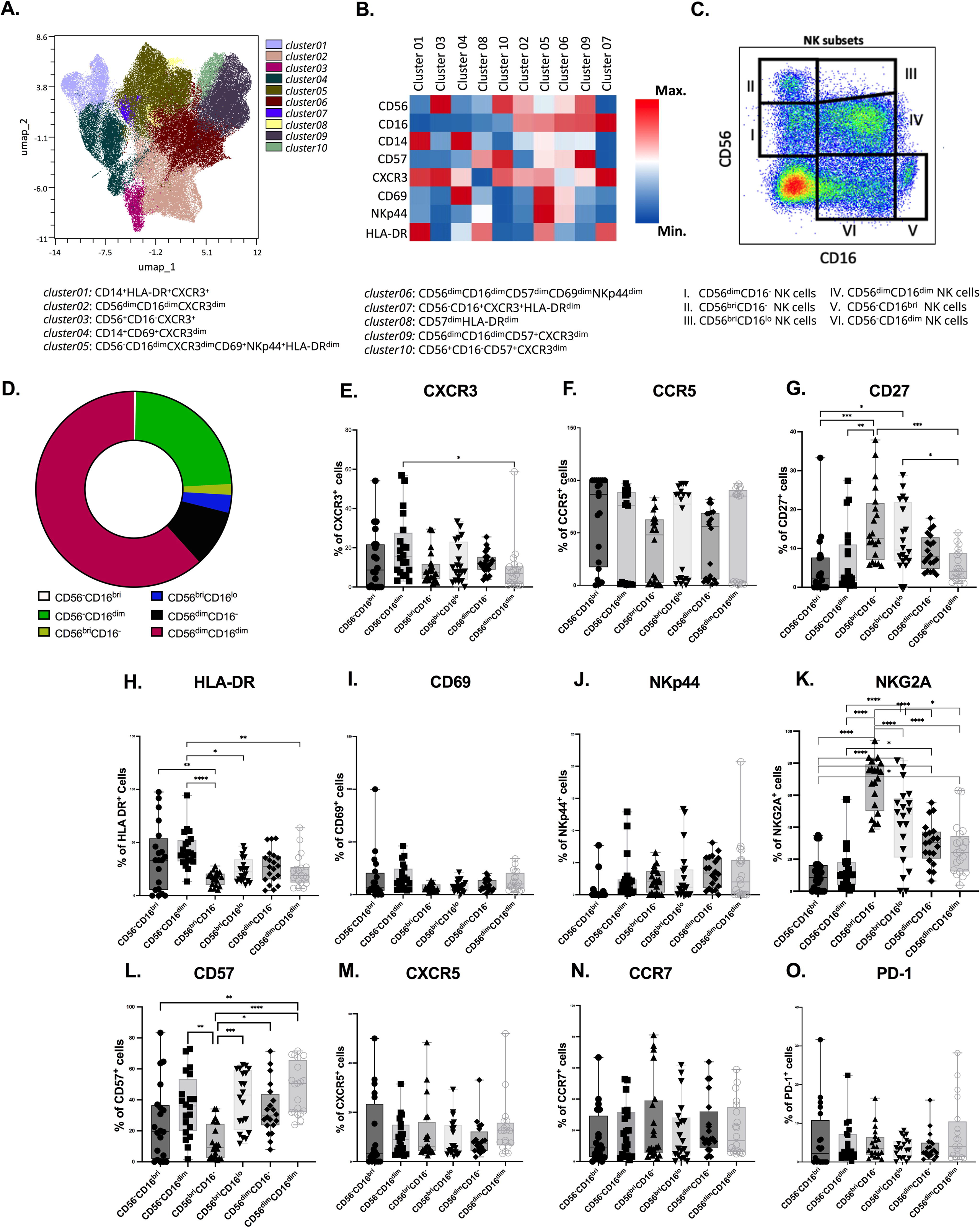

Additionally, we observed significantly increased CD27 expression (Figure1G) among CD56^bri^ NK subsets, specifically CD56^bri^CD16^−^ and CD56^bri^CD16^lo^ subsets, indicating their importance in mediating NK cell/T cell interaction via the CD27-CD70 pathway (37). CXCR3 expression (Figure 1E) was notably higher in the CD56^−^ CD16^dim^ population compared to other subsets suggesting that this subset has a greater potential for homing into inflamed tissue(38). The memory/maturity marker CD57 (Figure 1L) was elevated in the CD56^dim^CD16^dim^ cell subset. Importantly, we did not observe differences in CCR5, CXCR5, CCR7, CD69, NKp44 and PD-1 expression among different NK cell subsets in HIV-adolescents.

### Differential expression of immune markers on NK cell subsets in PHIV adolescents

Next, we compared NK cell phenotypes between PHIV adolescents and HIV-adolescents. Surprisingly, we did not discern any substantial differences in the relative representation of NK subsets between PHIV adolescents and their HIV-counterparts (Figure 2A). Nevertheless, when comparing the two groups, we did observe significant variations in the expression levels of multiple cell surface immune markers within NK cell subsets (Figure 2B-D). Significantly increased levels of surface receptors CD57, CXCR3, and NKp44 were observed within the CD56^dim^CD16^dim^ NK cell subset in PHIV adolescents compared to their healthy counterparts (Figure 2E-G). In contrast, expression of CD69 and HLA-DR were elevated on the CD56^dim^CD16^−^ NK cell subset in the PHIV group compared to expression on cells from the HIV-group (Figure 2H-I). These findings collectively indicate an overall activated phenotype of these NK cell subsets in PHIV. Notably, the CD56^dim^CD16^dim^ NK subset within the PHIV group exhibited numerous alterations in the surface expression of immune markers compared to other NK cell subsets (Supplemental Figure 2).

**Figure 2.**
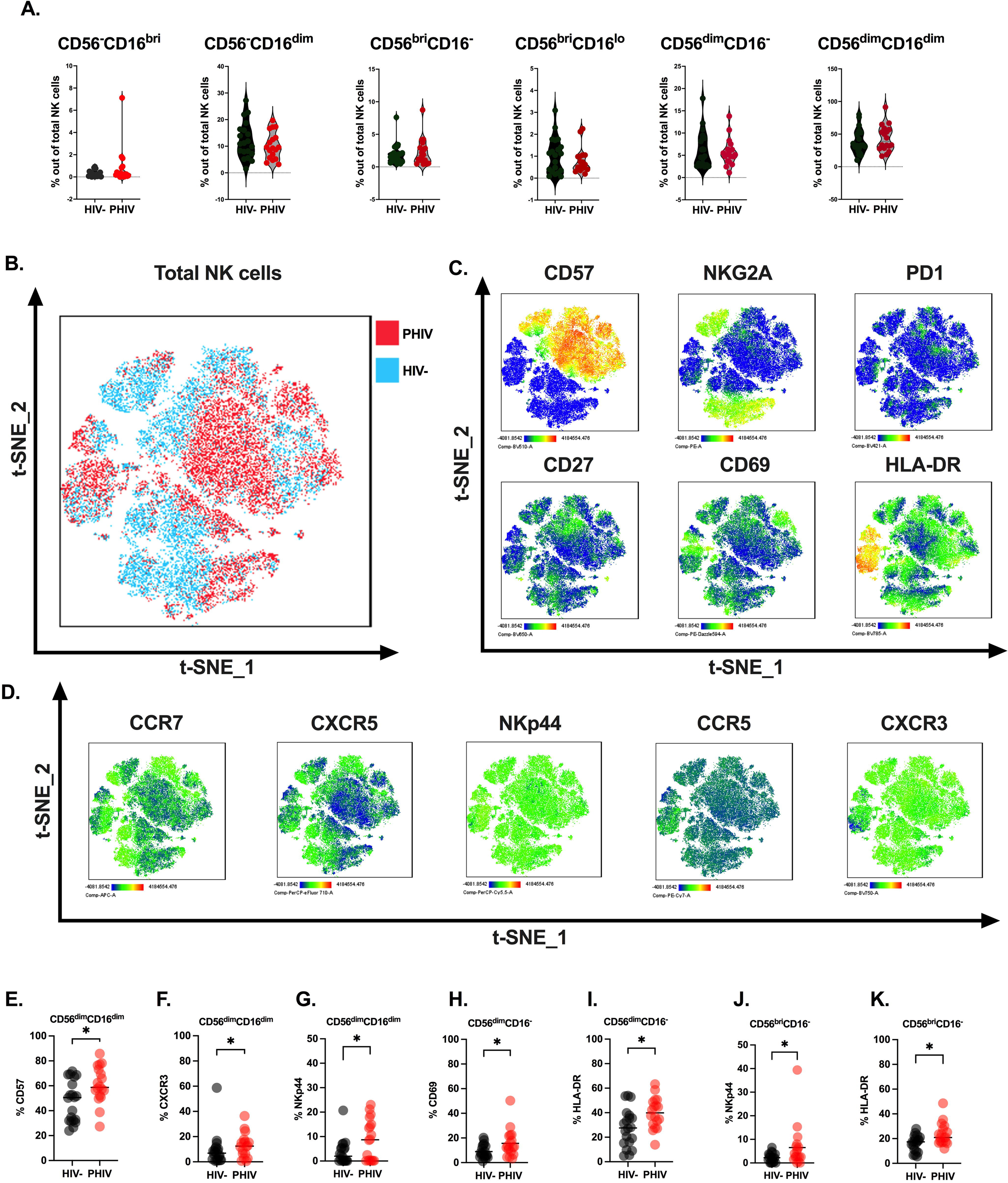

### Memory-like NK cell subset (CD57^+^) is highly present in PHIV adolescents compared to the HIV-group

CD57^+^ memory-like NK cells have emerged as a critically important component in the immune response to HIV. Furthermore, CD57 has been linked to cell adhesion and the acquisition of homing molecules, notably CXCR1 and CX3CR1, the ligands for which are predominantly expressed in inflamed peripheral tissues (39). In our study, we observed that among PHIV patients, the CD56^dim^CD16^dim^ NK cell subset exhibited higher levels of CD57 expression than other NK cell subsets (Figure 3A-C). Thus, we conducted a detailed analysis of the CD57^+^CD56^dim^CD16^dim^ population using conventional and high-dimensional data analysis approaches (Figure 3D-F). Our findings revealed that CXCR3 is markedly expressed in the CD57^+^CD56^dim^CD16^dim^ NK cell subset within the PHIV group when compared to levels in HIV^−^ adolescents (Figure 3G). This surprising finding suggest that in PHIV, memory-like NK cells (CD57^+^CD56^dim^CD16^dim^) could possess the ability to migrate to inflamed tissues.

**Figure 3.**
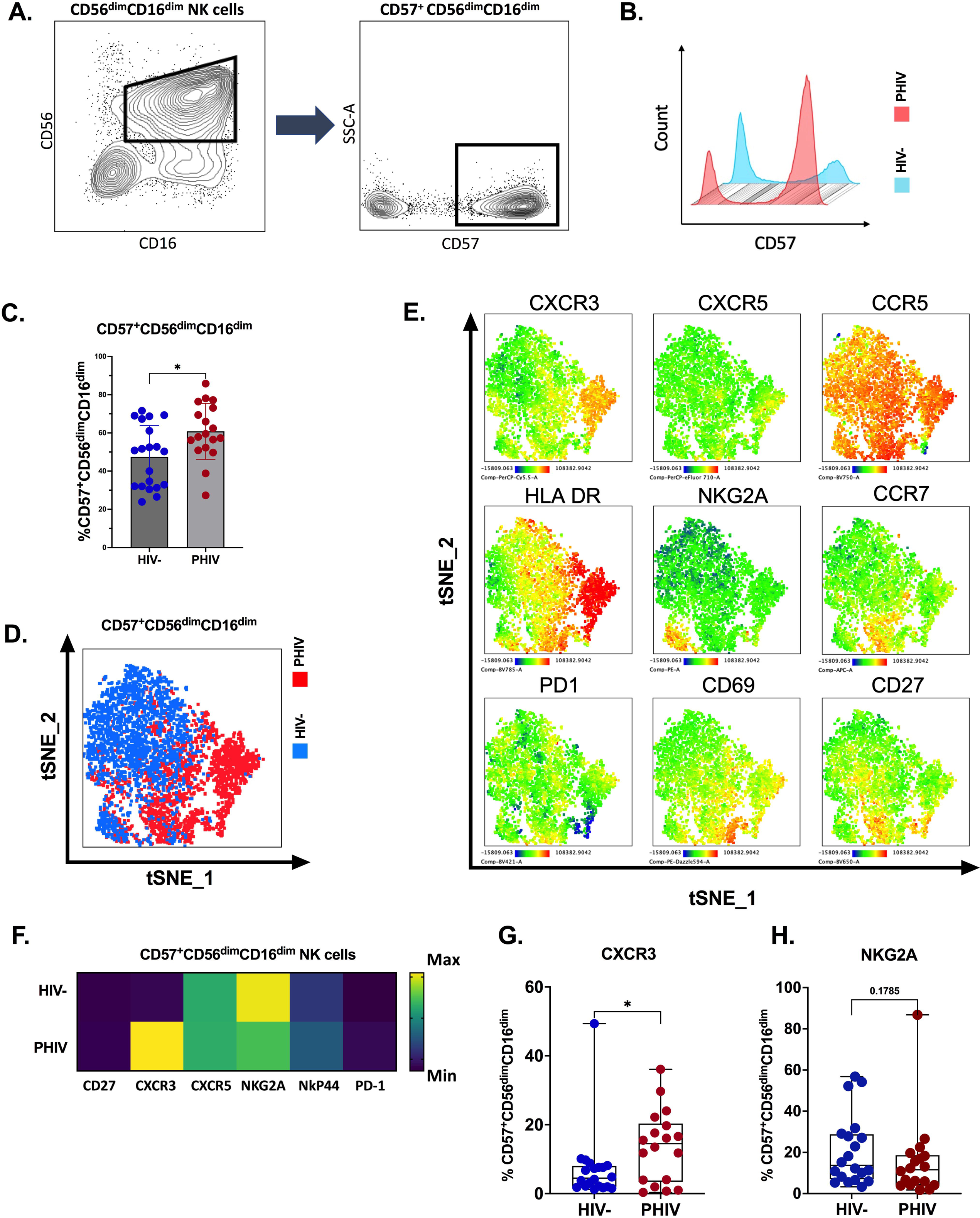

### Distinct relationships among cardiometabolic biomarkers, Ox-LDL, and NK cell subsets

Next, we measured plasma biomarkers of immune activation and inflammation among study participants. Although there were no statistically significant differences in the levels of many biomarkers between groups (Figure 4A and Table 3), surprisingly, the plasma concentration of Ox-LDL was reduced among PHIVs compared to the plasma concentration of Ox-LDL in adolescents without HIV (Figure 4B). Macrophages ingest Ox-LDL in the blood vessel wall, forming cholesterol-rich foam cells and initiating the development of atherosclerotic plaques. Pearson correlation coefficient analysis showed significant correlations between NK cell subsets and Ox-LDL in PHIV adolescents (Figure 4C). Notably, the memory-like NK cell subset (CD57^+^ CD56^dim^CD16^dim^), which was increased in PHIVs (Figure 4D), exhibited robust negative correlations with Ox-LDL, providing deeper insights into the complex relationship between memory-like NK cells and Ox-LDL. In addition, although the frequency did not change in the PHIV cohort, a strong negative correlation was observed between CCR5^+^ NK cell subsets (Figure 4E-H) and Ox-LDL levels, specifically within the PHIV cohort.

**Figure 4.**
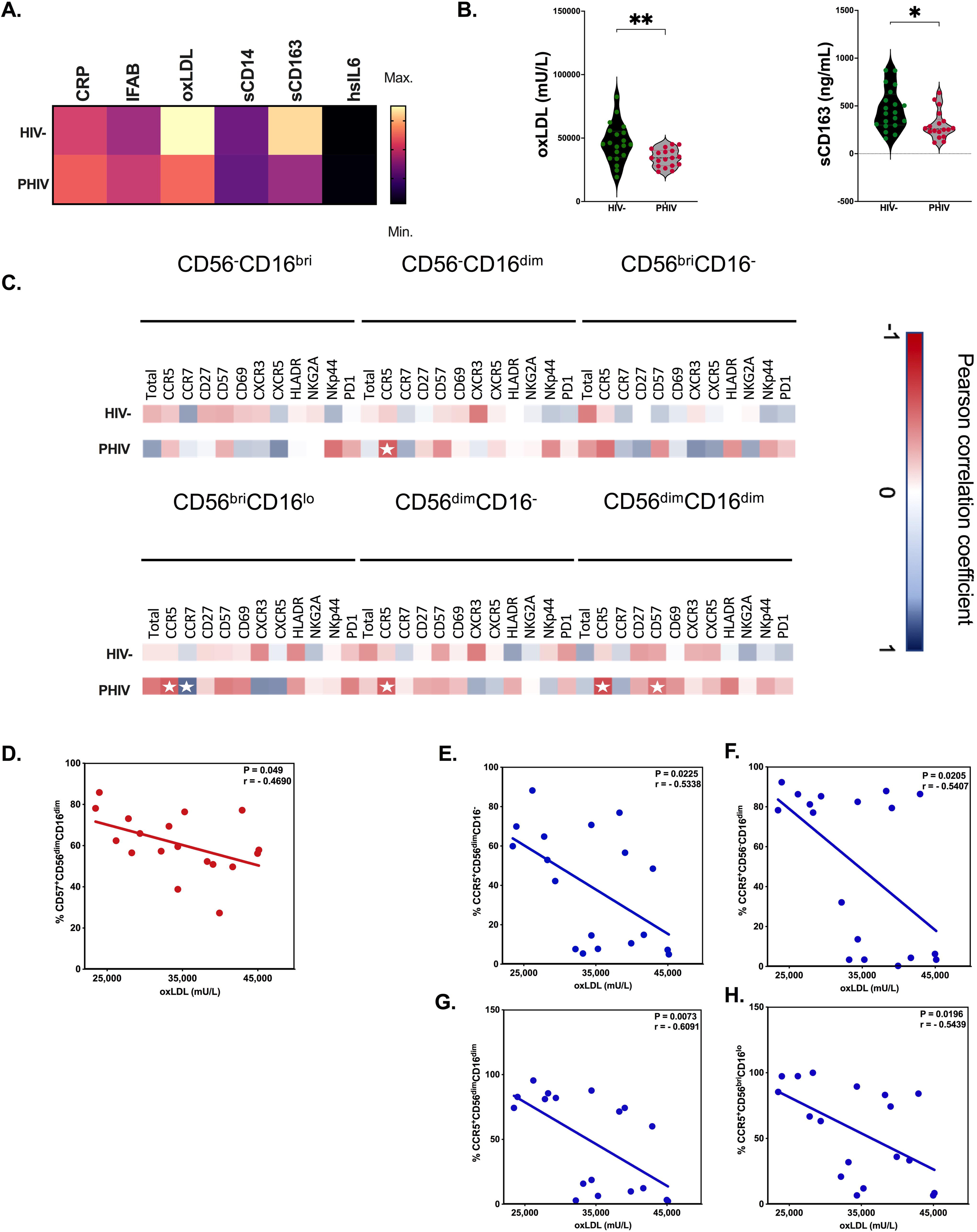

### Comprehensive transcriptomic profiling unveils differential regulation of innate immune and lipid metabolism pathways

We conducted an in-depth transcriptomic analysis and identified 1638 differentially expressed genes (DEGs) in PBMCs obtained from PHIV and HIV-adolescents. Among these statistically significant DEGs (p<0.05), 777 were upregulated, while 861 were downregulated (Figure 5A and Supplemental Figure 4A and B). In terms of innate immune activation, noteworthy upregulated DEGs included *MYD88*, *TLR1*, *TLR2*, *CLEC4D*, *CLEC12B*, *IRAK3, ITGB3BP*, *TNF*, and *CD163*, while the expression profiles of *GZMM* and *IL6R* were downregulated in PHIV. Additionally, we observed significant variations in gene expression associated with immune cell migration to inflamed tissues, including upregulation of *CXCL10*, *ITGB3BP*, and downregulation of *ITGB7*, *CCR4*, *CXCR6*, *CXCR5*, and *CCR6*. Our analysis also revealed altered transcriptional profiles related to vascular wall remodeling involving genes like *S1PR1*, *FLT4*, and *COL18A1*. Our flowcytometry analysis also revealed increased frequencies of intermediate monocytes (Supplemental Figure 3A) among PHIVs (p<0.05) and this was partially reflected through our transcriptome analysis that showed upregulated *CD36* and *CLEC10A*, which are known to be increasingly expressed in this subset of monocytes (40, 41). *CD36* encodes for the Ox-LDL receptor itself while *COLEC12* (also found to be upregulated among PHIVs) mediates the binding and internalization of Ox-LDL. Furthermore, our analysis predicted several epigenetic modifications occurring in PBMCs of PHIV adolescents. This prediction was substantiated by the high number of DEGs associated with histone modification and DNA methylation, exemplified by genes like *HIST1H1C*, *HIST1H2AH*, and *HDAC1*. We conducted functional and pathway enrichment analyses to ascertain the biological significance of these DEGs. Our gene ontology analysis revealed a profound impact on vascular and lipid metabolism-related biological processes involving immune cells among PHIV, including ‘leukocyte adhesion to endothelial cells’, ‘NK cell activation’, ‘NK cell degranulation’, ‘myeloid cell migration’, ‘myeloid cell differentiation’, ‘regulation of lipid storage’, ‘regulation of lipid catabolism’, and ‘vascular wall remodeling’ (Figure 5B and supplemental Figure 5). Overall, the protein-protein interaction network analysis highlighted the enrichment of pathways specifically related to innate immunity and epigenetic regulation (Figure 5C). Observed clustering based on the top differentially expressed genes and transcriptional pathways (Supplemental Figure 6 A and B), underscores the phenotypical and functional distinctions that exist between PHIV adolescents compared to HIV unexposed controls, offering insights into the intricate transcriptional profiles shaping these differences.

**Figure 5.**
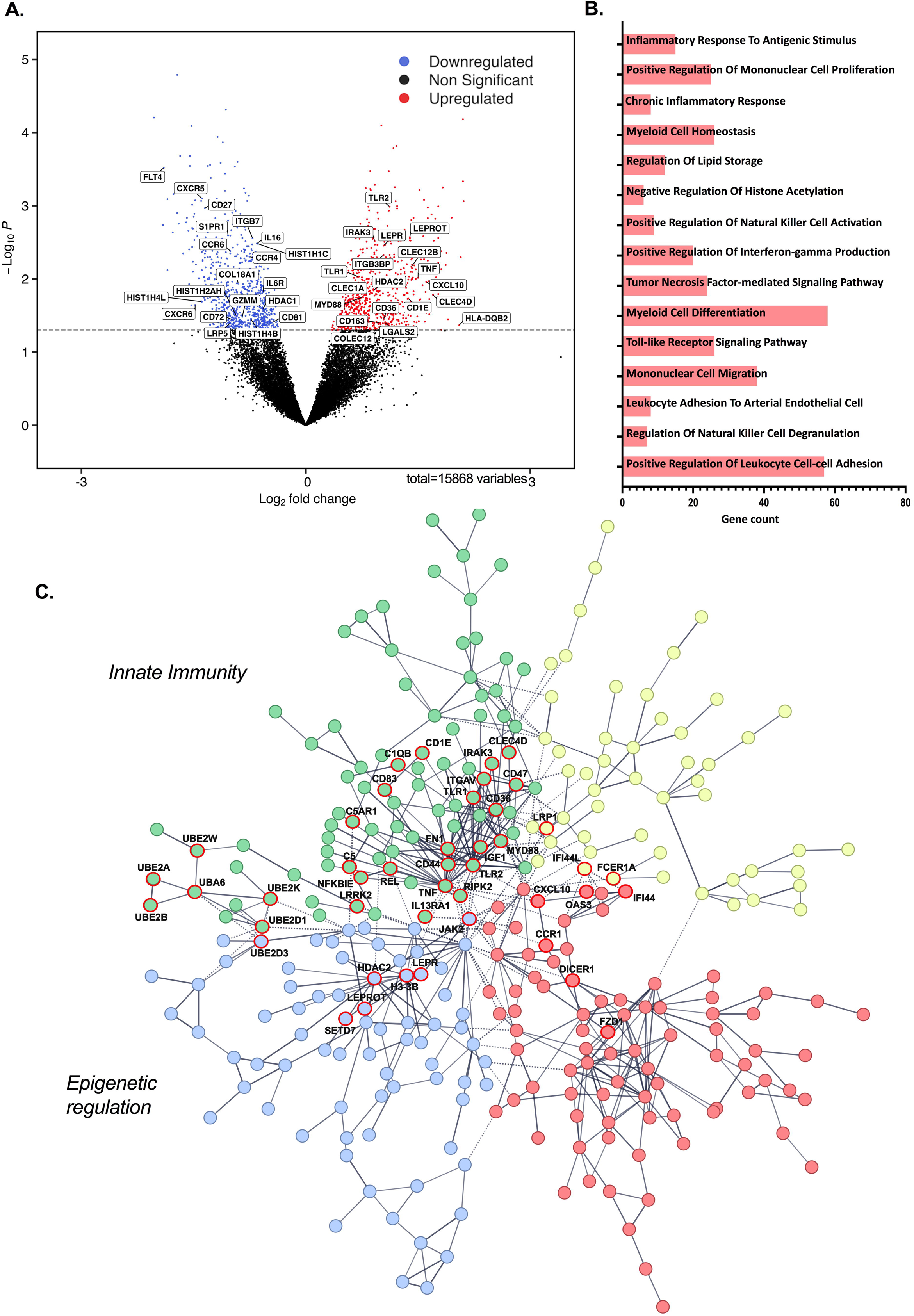

### Activated NK cell-mediated enhanced Ox-LDL uptake by monocyte-derived macrophages in-vitro

Based on our comprehensive analysis, we posit that in adolescents with PHIV, activated NK cells, especially those expressing CCR5 or exhibiting memory-like characteristics, interact with macrophages derived from blood monocytes, leading to enhanced uptake of Ox-LDL. It has been documented that in established atherosclerosis, plasma levels of Ox-LDL correlate with levels within established and saturated plaques (42). However, our results from an adolescent HIV population with no known risk factors for atherosclerosis and less likelihood of plaques suggest increased uptake of Ox-LDL by monocyte-derived macrophages from the bloodstream concurrently reduces plasma Ox-LDL levels, the initiation of atherogenesis. We conducted an in vitro experiment to examine the influence of activated NK cells on the uptake of Ox-LDL by macrophages (Figure 6A). We isolated NK cells from PBMCs of HIV-donors (n=5). These NK cells were externally activated using IL-12, IL-15, and IL-18 (43). Concurrently, we induced the formation of autologous monocyte-derived macrophages (MDMs) from peripheral blood monocytes by allowing PBMCs to rest in Teflon-coated wells for up to six days, as described previously (44). After six days, we incubated these MDMs in a medium containing commercially prepared Dil-labeled Ox-LDL for four hours, in the presence and absence of activated autologous NK cells. We employed spectral flow cytometry analysis to measure the uptake of Dil-Ox-LDL. In all donor samples tested, we observed significantly higher Dil-Ox-LDL uptake in the presence of activated NK cells (Wilcoxon signed-rank test; p<0.001 for all) (Figure 6B and C). To further investigate the role of activated NK cells in Ox-LDL uptake, we introduced blocking antibodies against NKG2D while co-culturing NK cells with monocyte-derived macrophages (MDMs). This experiment revealed significantly reduced Ox-LDL uptake in all tested samples in the presence of NKG2D blocking (p<0.001). These findings confirm the role of activated NK cells in enhancing Ox-LDL uptake by MDMs and also suggest that the NK cell receptor NKG2D may play an essential role in this process (Figure 6B and C). This adds to previous reports of HIV-associated increased uptake of Ox-LDL in to MDMs associated with high TNF-α levels, where TNF-α upregulates the NKG2D ligand, MICA, in the intimal tissue environment (45, 46).

**Figure 6.**
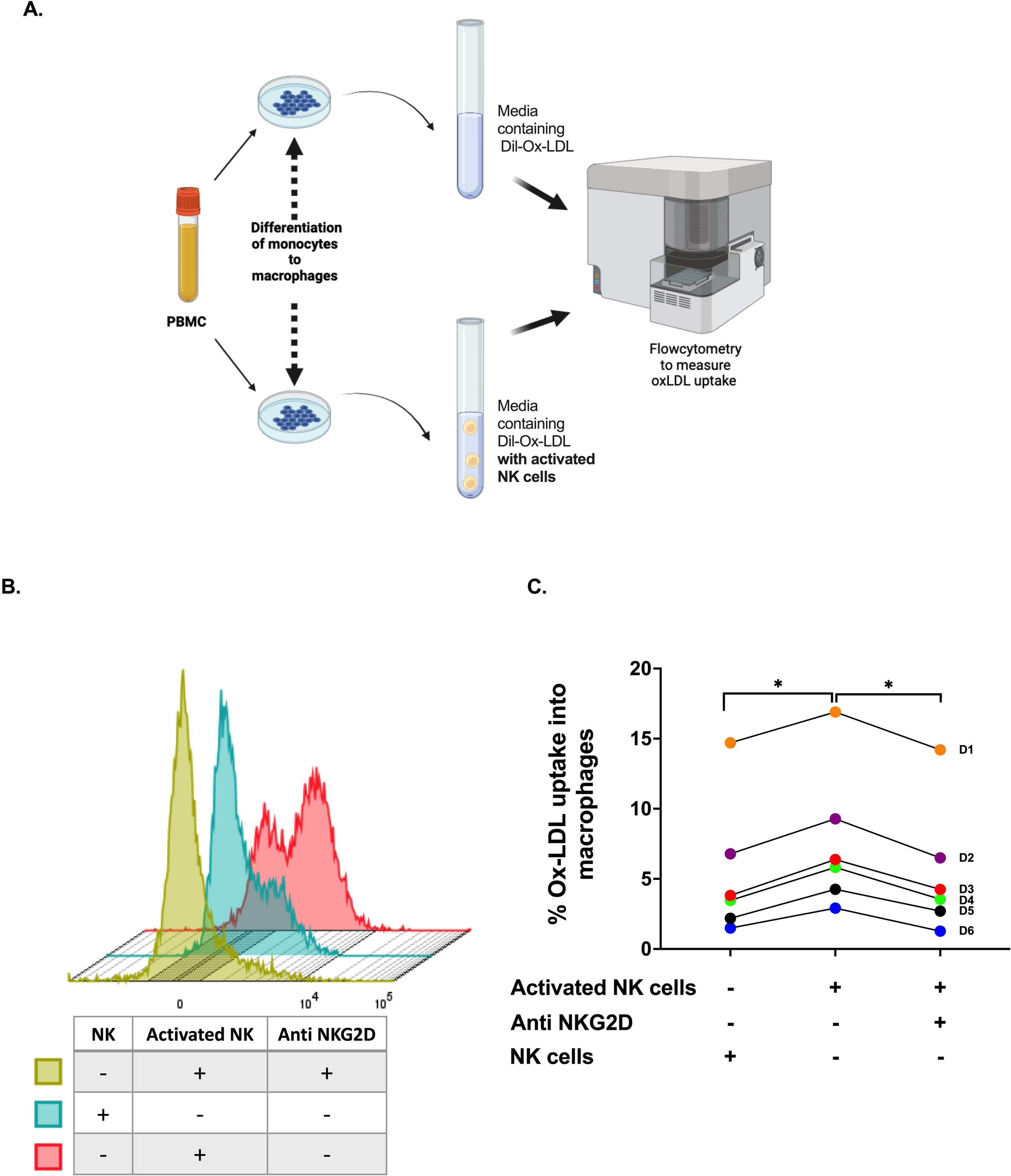

## Methods

### Study participants

This study took placed at the Joint Clinical Research Center (JCRC) in Kampala, Uganda between 2017-2021 as previously described (32). Participants (PHIV and HIV-children) were prospectively enrolled in this observational cohort. All participants were 10-18 years of age and as per Ugandan research guidelines, provided written informed assent. By design, PHIV participants (n=18; median age =14.4 years) were on ART for at least 2 years with a stable regimen for at least the last 6 months with HIV-1 RNA < 50 copies/mL. Self-reported co-infections and malnutrition were exclusionary. Adolescents with pregnancy or intent to become pregnant were excluded. The control group consisted of age/sex matched HIV-uninfected adolescents from the same geographical location (n=20; median age=14.2 years) who had not been exposed to HIV in utero. None of the participants were suffering from acute infection, malnutrition, were pregnant, had active tuberculosis or diabetes mellitus, or were on anti-inflammatory or cholesterol lowering medication at the time of sample collections.

### Cell collection and processing

Participants were seen for blood draw obtained after an 8 hour fast. Plasma, serum, and PBMCs were cryopreserved and shipped to University Hospitals Cleveland Medical Center, Cleveland, Ohio. All assays were performed in batches and without prior thaw. For the purpose of this analysis, we included participants with plasma, serum and PBMCs available at the same time point.

### Flow cytometry cell staining and analysis

We assessed innate and adaptive immune cell surface markers in PHIV adolescents and HIV-uninfected controls. Cryopreserved PBMCs were resuspended at 1–2 million cells per ml in R10 media (RPMI-1640 supplemented with 10% FBS [fetal bovine serum], 2 mM l-glutamine, 100 U/mL penicillin, and 100 mg/mL streptomycin) and surface staining for flowcytometry was performed. Samples were stained with Fixability Viability Dye (Zombie NIR Cat No. 423105) and incubated for 15 minutes. Cells were then washed, followed by the addition of a surface antibody cocktail, the antibodies being previously titrated to determine the optimal concentration (antibodies used in this study are listed in Table 4). After a 30-minute incubation period cells were washed once again (All washing steps were carried out at 700g for 5 min at 4 °C). Following the filtering of cells through strainer capped FACS tubes, samples were acquired on a Cytek Aurora flow cytometer. Spectral flow cytometry data were analyzed using FlowJo software (Tree Star). We obtained “fluorescence minus 1” controls (cells stained with all fluorochromes used in the experiment except 1) for each marker prior to spectral unmixing (Supplemental Figure 1B).

### Measurement of plasma biomarkers

The plasma collected from blood samples were stored at –80^0^C and batched until processing without a prior thaw. Then, using an enzyme-linked immunosorbent assay (ELISA), we measured the following inflammatory biomarkers in the plasma of PHIV adolescents and uninfected controls: soluble CD14 (sCD14), C reactive protein (CRP), oxidized low-density lipoprotein (Ox-LDL), 1,3-β-D glucan (BDG), soluble CD163 (sCD163), intestinal fatty acid binding protein (IFAB) and high-sensitivity interleukin-6 (hsIL6) (47).

### Invitro assay for Ox-LDL uptake by monocyte-derived macrophages (MDMs)

We conducted an in vitro experiment to investigate the impact of activated NK cells on the uptake of Ox-LDL by MDMs to substantiate our in vivo observations. Venous blood samples were collected in heparinized tubes in order to isolate PBMCs via Ficoll gradient centrifugation. NK cells were then isolated from these PBMCs through negative selection using the Miltenyi Biotec NK cell isolation kit (130-092-657). Isolated NK cells were incubated invitro with IL-12, IL-15 and IL-18. Additional PBMCs isolated from the same donor were rested and incubated in Teflon-coated wells for up to six days, to induce differentiation of monocytes into macrophages (MDMs) (44). After six days, we introduced these MDMs to a medium containing commercially manufactured Ox-LDL that had been tagged with Dil dye to measure its uptake (10 ug/mL) into MDMs during a four-hour incubation period both in the presence and absence of activated NK cells. Cells were washed twice with PBS, and the intensity of DiI staining was measured by flow cytometry. To identify MDMs, CD45, CD3, CD19, and CD14 surface immune markers were used. The above experiment was also carried out in the presence of NKG2D and IFNγ blocking antibodies (added during the four-hour incubation step with along with activated NK cells) on separate occasions to investigate the mechanism of NK cell-influence on MDMs to impact their uptake of Ox-LDL.

### RNA Extraction and QC

Cell viability was measured prior to RNA extraction for PBMC samples using the Countess II FL Automated Cell Counter (ThermoFisher), and RNA extraction was performed using the RNeasy mini kit (Qiagen). The quality of all RNA was determined using the RNA integrity number as calculated from the Agilent Fragment Analyzer with the SS RNA assay kit (Agilent) while the concentration was determined by NanoDrop (ThermoFisher).

### Bulk RNA Sequencing

Total RNA prepared above was normalized to 100 ng input for library preparation with the TruSeq Stranded Total RNA with Ribo-Zero Globin kit (Illumina). The resulting libraries were assessed on the Agilent Fragment Analyzer with the HS NGS assay (Agilent) and quantified using the NEBNext® Library Quant Kit for Illumina® (New England Biolabs, Inc.) on an Applied Biosystems QuantStudio 7 Flex Real-Time PCR (ThermoFisher). Medium depth sequencing (30 million reads per sample) was performed with Novaseq (Illumina) on one SP flow cell as a 100 cycle paired-end run.

### RNA-seq analysis

Raw demultiplexed fastq paired end read files were trimmed of adapters and filtered using the program skewer (48) to throw out any with an average phred quality score of less than 30 or a length of less than 10. Trimmed reads were then aligned using the STAR (49) aligner to the Homo sapiens NCBI reference genome assembly version GRCh38 and sorted using SAMtools (50). Aligned reads were counted and assigned to gene meta-features using the program featureCounts (51) as part of the Subread package. These count files were imported into the R programming language and were assessed for quality control, normalized and analyzed using an in-house pipeline utilizing the limma voom method with quantile normalization (52) for differential gene expression analysis. Gene set variation analysis was performed using the GSVA Bioconductor library and the Molecular Signatures Database v5.0 (53, 54). Protein-protein interaction visualizations were generated using Stringdb (55). Gene Ontology (GO) enrichment analysis was performed to gain insights into the biological processes associated with a set of genes of interest. The analysis was carried out using the clusterProfiler package in R (version 4.2.1). The significance threshold for enriched GO terms was set at a p-value cutoff of 0.05, and the Benjamini-Hochberg method was applied for multiple testing correction using GO terms with a q-value (adjusted p-value) below 0.05 were considered statistically significant. Cytoscape software (version 3.10.0) was used to visualize significant GO terms (56).

### Statistical Analyses

Graphs and heatmaps were prepared using Graph Pad Prism (version 9.3.1), while dot plots, contour plots and t-SNE dimensionality reduction plots were generated using FlowJo (version 10.8.2). Data were statistically analyzed using their bundled software. Statistical comparisons involving more than two experimental groups were performed using ANOVA followed by Tukey’s multiple comparison test for pairwise comparisons. Comparisons between two groups were performed using the Wilcoxon rank-sum test. To analyze the relationship between immune signatures and plasma biomarkers, the Pearson correlation coefficient was calculated and plotted on a correlogram using R 4.2.1. Scatter plots showing linear relationships between data were created for significant correlations.

### Study Approval

We obtained approval by the Research Ethics Committee in Uganda, University Hospitals Cleveland Medical Center, Cleveland, Ohio and The Ohio State University. Caregivers gave written informed consent.

## Discussion

In this study we performed a comprehensive analysis of the major innate immune cells (i.e., NK cells and monocytes) in a group of PHIV adolescents and HIV-age/sex matched controls in Uganda. Our results demonstrated that several subsets of NK cells are activated (as evidenced by their expression of CD69, CD57, NKp44 and HLA DR) in our PHIV cohort (Figure 2). Furthermore, in the PHIV cohort, subsets including CD56^dim^CD16^dim^ NK cells had increased levels of expression of CXCR3, granting them a greater potential to migrate to inflamed tissue, compared to those in the HIV-uninfected adolescents. Our correlational analyses (Figure 4C-H) revealed a negative association between plasma levels of Ox-LDL and many NK subsets expressing CD57 and the chemokine receptor CCR5, suggesting their involvement in the lowering of Ox-LDL in the plasma compartment. This was confirmed by bulk RNA sequencing data of PBMCs which revealed the enrichment of several DEGs and transcriptional pathways related to NK cell activation, innate immune activation, and Ox-LDL metabolism. The direct influence of active NK cells in promoting uptake of Ox-LDL by tissue macrophages was further supported by our in vitro studies (Figure 6). NK cells have traditionally been considered as important mediators of anti-tumor(57) and anti-viral (58) immunity. More recently, they have been also linked to various pathophysiological disease processes including cardiovascular diseases(59). NK cells are known to be key players in atherosclerosis(31), the most common pathophysiological condition associated with ischemic heart disease. Most of the evidence on the involvement of NK cells in atherosclerosis has been derived from observational studies conducted on plaque tissue using immunohistochemistry. Bonaccorsi et al. studied phenotypic and functional characteristics of NK cells within the atherosclerotic plaque environment and identified increased frequencies of CD56^bri^perforin^lo^ NK cells, expressing tissue-resident markers such as CD103, CD69 and CD49a, and producing increased amounts of IFNγ(60).

Chronic HIV infection results in a persistent pro-inflammatory state in people living with HIV, even during viral suppression by ART. Similar to previous studies(61), we too observed increased activation among several NK subsets in the PHIV cohort. These cells had increased expression of activation markers such as CD69, NKp44 and HLA-DR in many NK subpopulations, and bulk RNA-seq data confirmed upregulated transcription of signaling adaptors like MyD88 which is critical for NK and other immune cell activation(62) along with enriched GO terms like ‘NK cell activation involved in immune response’.

We hypothesize that expression of the chemokine receptor CCR5 in these activated NK subsets in the PHIV population may signify a much greater potential of these cells to influence the pathogenesis of cardiometabolic disease. Compared to HIV-individuals, PLWH have been found to demonstrate significant arterial wall inflammation (63). During this inflammation, activated dendritic cells within vascular tissue secrete various chemokines such as RANTES/CCL5, CCL4, CX3CL1, CXCL10(64), and this has been supported by our transcriptome data. Particularly in the case of vascular inflammation surrounding atherogenesis, murine studies have demonstrated an abundance of CCL5 and a high proportion of the chemokine receptor CCR5-expressing immune cells in the atherogenic plaque microenvironment (65). Several activated and mature NK subsets were observed among PHIV adolescents (as evidenced by their increased expression of activation and maturation markers) that are pro-inflammatory in nature and express the chemokine receptor CCR5 can potentially migrate towards vascular tissue that express the corresponding chemokine CCL5 (66). It is likely that vascular tissue-homing activated, pro-inflammatory NK cells likely express high levels of additional activating receptors like NKG2D on their surface (67). As highlighted by our in vitro experiments, these cells may engage in NKG2D-mediated crosstalk with vascular tissue-resident macrophages to promote their increased uptake of plasma Ox-LDL. The strong negative correlation we observed between several of these increasingly activated NK subsets among PHIVs expressing CCR5 and plasma Ox-LDL thus suggest a potential mechanism that may contribute to the low plasma Ox-LDL levels observed among our PHIV cohort. This may potentially result in the increased likelihood of atherogenesis among these individuals as opposed to HIV-individuals.

Migration of blood monocytes into the blood vessel wall and the subsequent differentiation of these cells into macrophages likely drives early stages of atherosclerotic plaque development (68). Among these, CD14^+^CD16^+^ intermediate monocytes have been found to be the key monocyte subset involved in atherogenesis (69). Justo-Junior et al. found that individuals with unstable angina had higher numbers of intermediate monocytes in the circulation and that these expressed high amounts of CCR2 which would mediate migration of these cells to inflamed vascular tissue expressing their ligand MCP-1(70). Interestingly, we also found a significant increase in the levels of circulating intermediate monocytes among our PHIV adolescents (Supplemental Figure 3), along with the enriched GO terms involving mononuclear cell proliferation, differentiation and migration, and we can only assume that the stage is slowly being set for the development and progression of a pro-atherogenic state in these individuals beginning very early on in their lives.

Results from our in vitro studies indicate that NK cells influence uptake of Ox-LDL by macrophages that in HIV, are more likely to migrate to inflamed vessel walls. Upon internalization of Ox-LDL using scavenger receptors such as CD36, LOX-1 and SR-A, macrophages differentiate into foam cells (71). The hypothesis that these macrophages actively interact with NK cells in the atherosclerotic lesion has been under investigation, and is supported by the fact that ligands like ‘major histocompatibility complex class I chain-related (MIC)’ (67) to the NK cell-activating receptor, NKG2D, have been found to be expressed on foam cells derived from macrophages exposed to Ox-LDL(72). Murine studies have shown that preventing NKG2D/ligand interaction suppresses plaque formation in ApoE2/2 mice(73). However, since NKG2D has also been found on subsets of T lymphocytes(67), T cells may also contribute to atherosclerotic plaque formation in this manner (as mentioned above). Our in vitro NKG2D blocking studies clearly demonstrate that NKG2D-mediated interactions between activated NK cells and macrophages increases macrophage uptake of Ox-LDL. This further supports our correlational analysis between plasma Ox-LDL levels and frequencies of activated NK cell subsets with migratory potential in our PHIV adolescents. Interestingly we also reported upregulated transcription of *CD36* in our PHIV cohort, which encodes for the receptor mediating uptake of Ox-LDL into macrophages, further promoting plaque generation. Indeed, in adults with HIV we have demonstrated increased expression of CD36 in monocytes, suggesting that numerous mechanisms collectively contribute towards a proatherogenic state among PHIVs (74).

It has been found that endogenous metabolites, such as Ox-LDL, that are abundant in the plaque microenvironment, train the generation of pro-atherogenic monocytes and macrophages (25). Dominant NK cell subsets in our PHIV cohort expressing high levels of memory markers like CD57(39) indicates that exposure of these cells to a lipoprotein-rich microenvironment may trigger trained immunity in these NK cells through epigenetic modification, similar to that seen in pro-atherogenic macrophages. Results from our bulk RNA-seq clearly demonstrate some degree of epigenetic modifications occurring in the leukocytes of PHIV adolescents. In the case of adolescents with HIV with no other dominant risk factors for cardiovascular disease, these trained NK cells (either by persistent viral replication, gut microbial translocation, co-pathogens, ART and/or Ox-LDL) may be considered as key innate immune cells involved in the ongoing progression of atherogenesis, by interacting with pro-atherogenic macrophages.

In conclusion, we show that in PHIV adolescents, activated, pro-inflammatory NK cell phenotypes associated with HIV infection may directly influence the uptake of circulating Ox-LDL by vessel wall-resident macrophages and contribute to the development of atherogenesis. This is concerning and may lead to increased risk of atherosclerotic CVD as these adolescents reach adulthood.

Furthermore, identification of the importance of the NKG2D/ligand interaction between NK cells and macrophages in atherogenesis may represent valuable novel targets for both therapeutic and prophylactic measures in mitigating cardiovascular disease in this patient population.

## Acknowledgments

The authors gratefully acknowledge the support of all participants and their families. We thank Wendy Lin and the staff of the CWRU Applied Functional Genomics Core for their technical assistance with the RNA sequencing. This work was supported by National Institutes of Health (NIH/NIAID) grant U01 AI168630-01.

## Data Availability

The data supporting the findings of this study are available upon request from the corresponding author. Data sharing is subject to ethical considerations and compliance with institutional policies.

## Author Contribution

M.A, M.G. performed experiments, data analysis and wrote the manuscript, A.K, K.A, performed experiments, V.M. C.K performed sample collections, B.R., B.T., M.R., W.L performed experiments, transcriptome data analysis, D.K., supervised data analysis, T.D. provided intellectual input and study design, C.C. M.C performed transcriptome data analysis, S.D.F, N.F and N.L designed the study and wrote the manuscript.

