## Supplementary figures and images for "Activated NK Cells with Pro-inflammatory Features are Associated with Atherogenesis in Perinatally HIV-Acquired Adolescents"

### Supplemental Figure 1

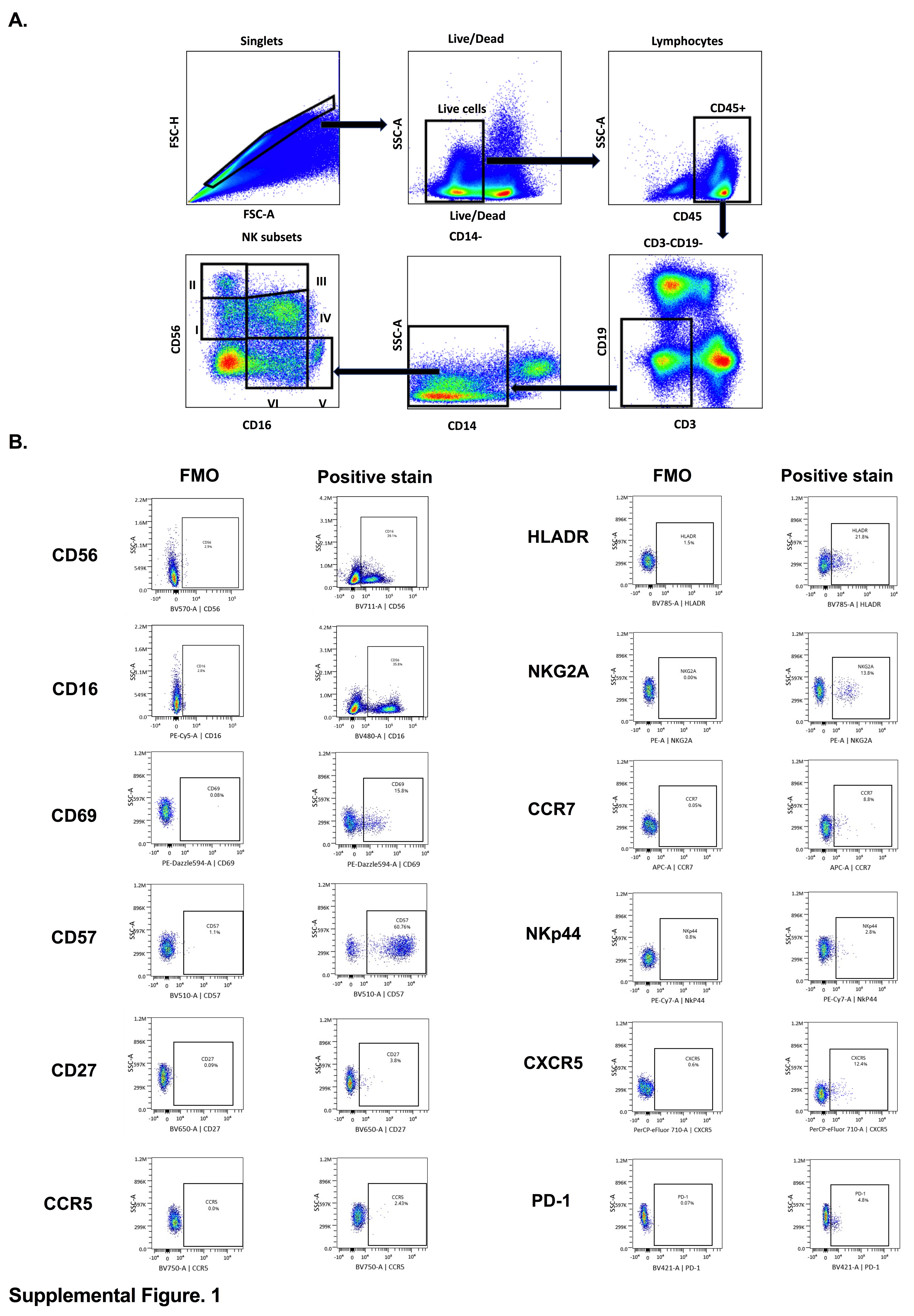

### Supplemental Figure 2

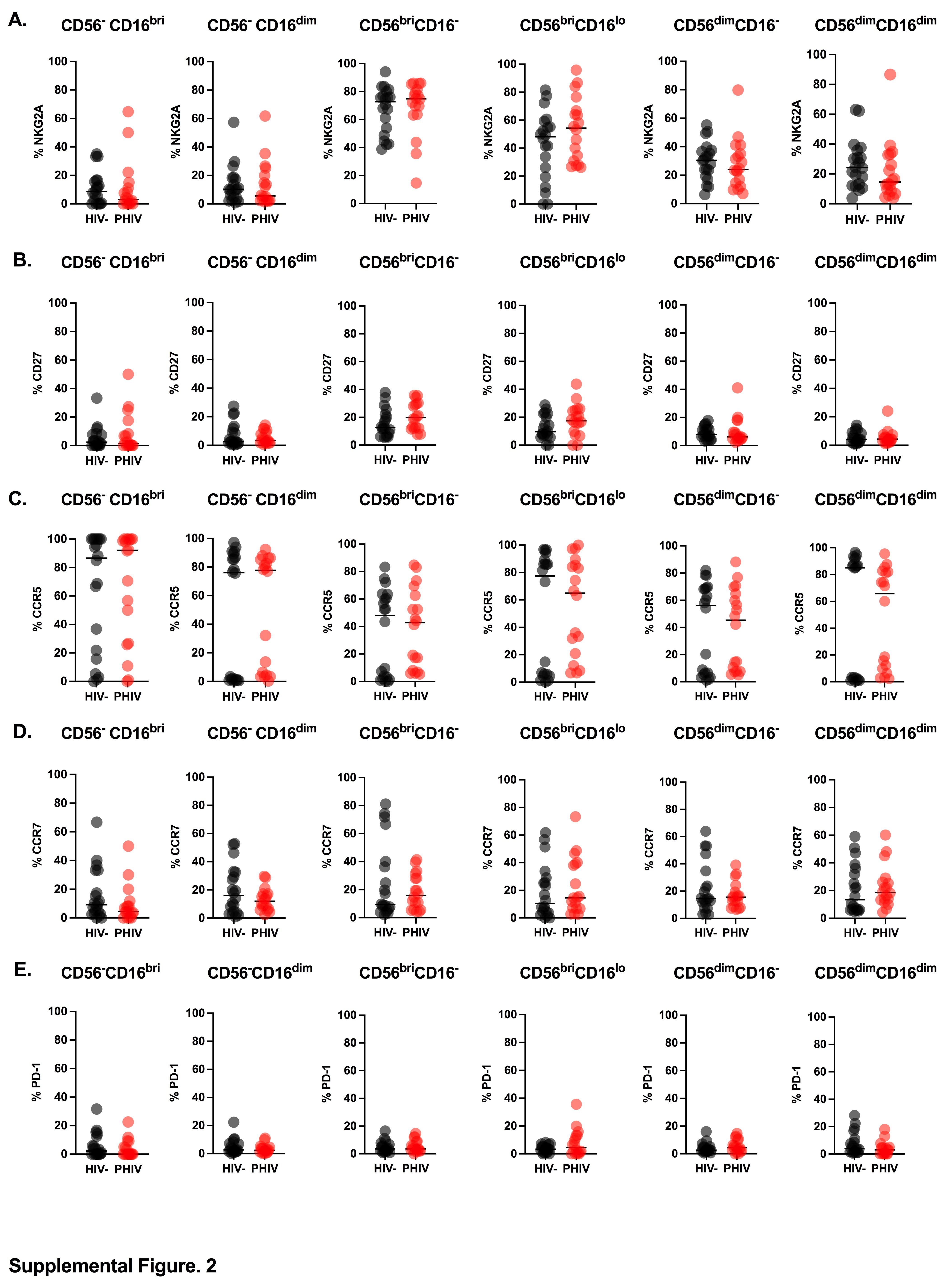

### Supplemental Figure 3

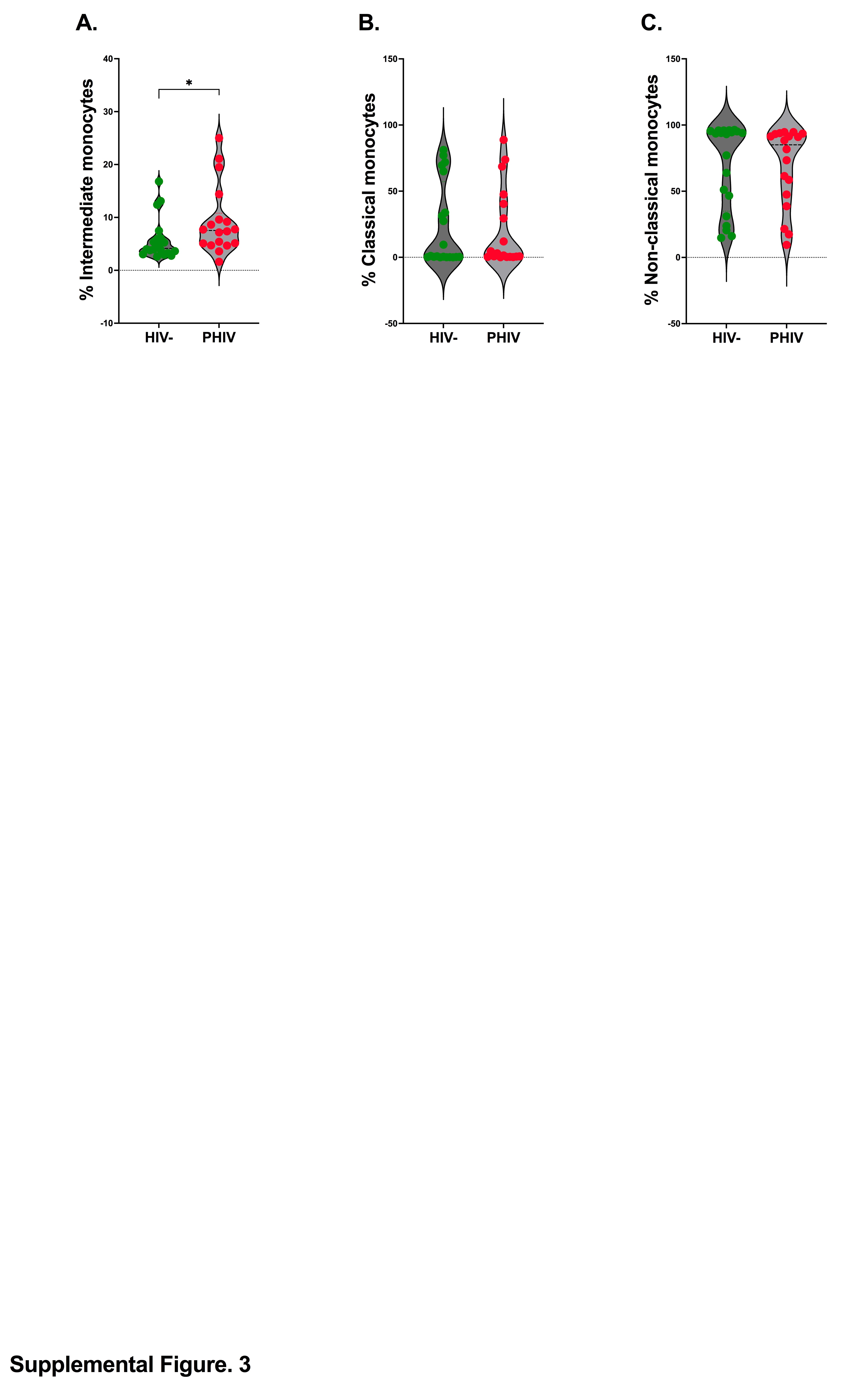

### Supplemental Figure 4

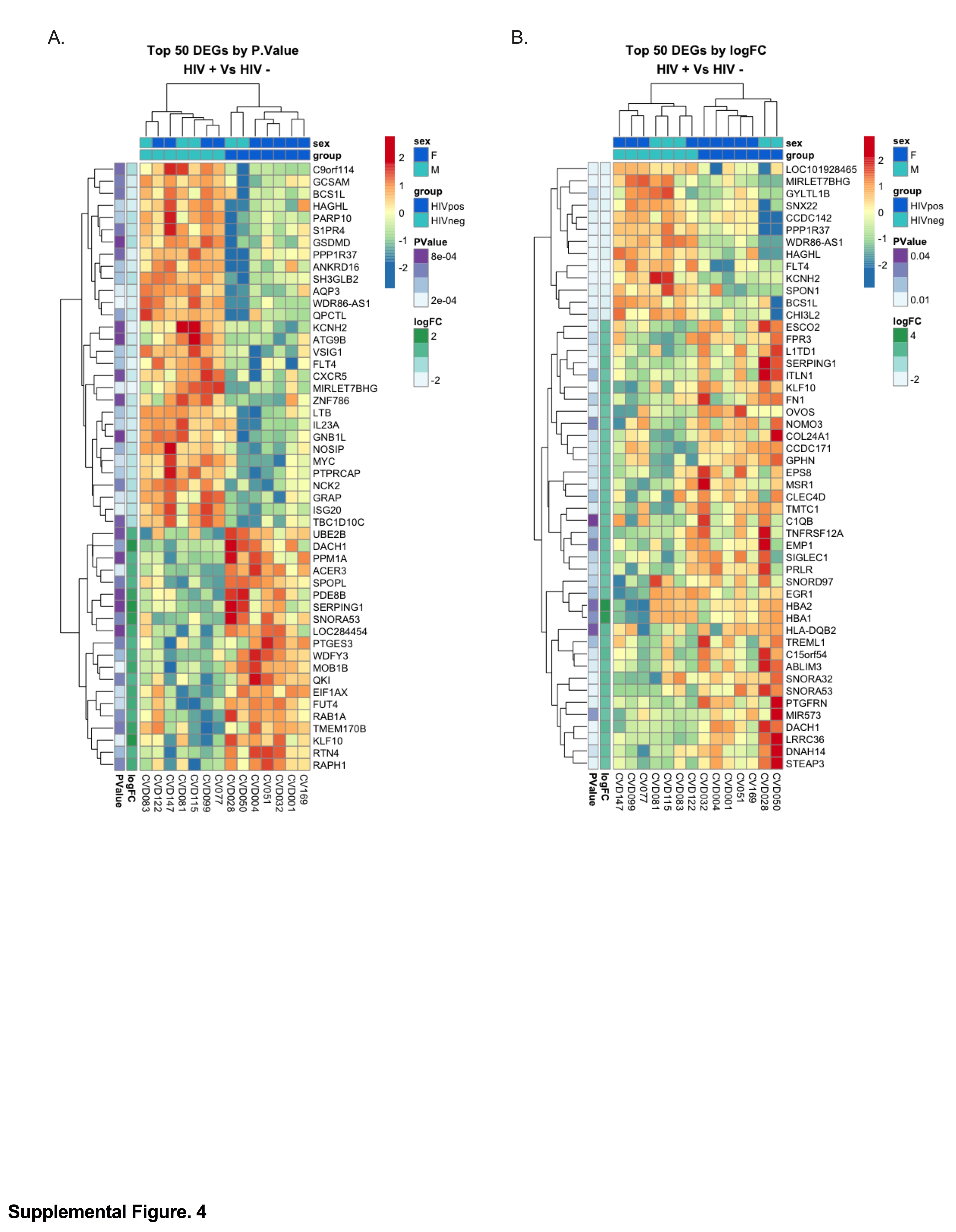

### Supplemental Figure 5

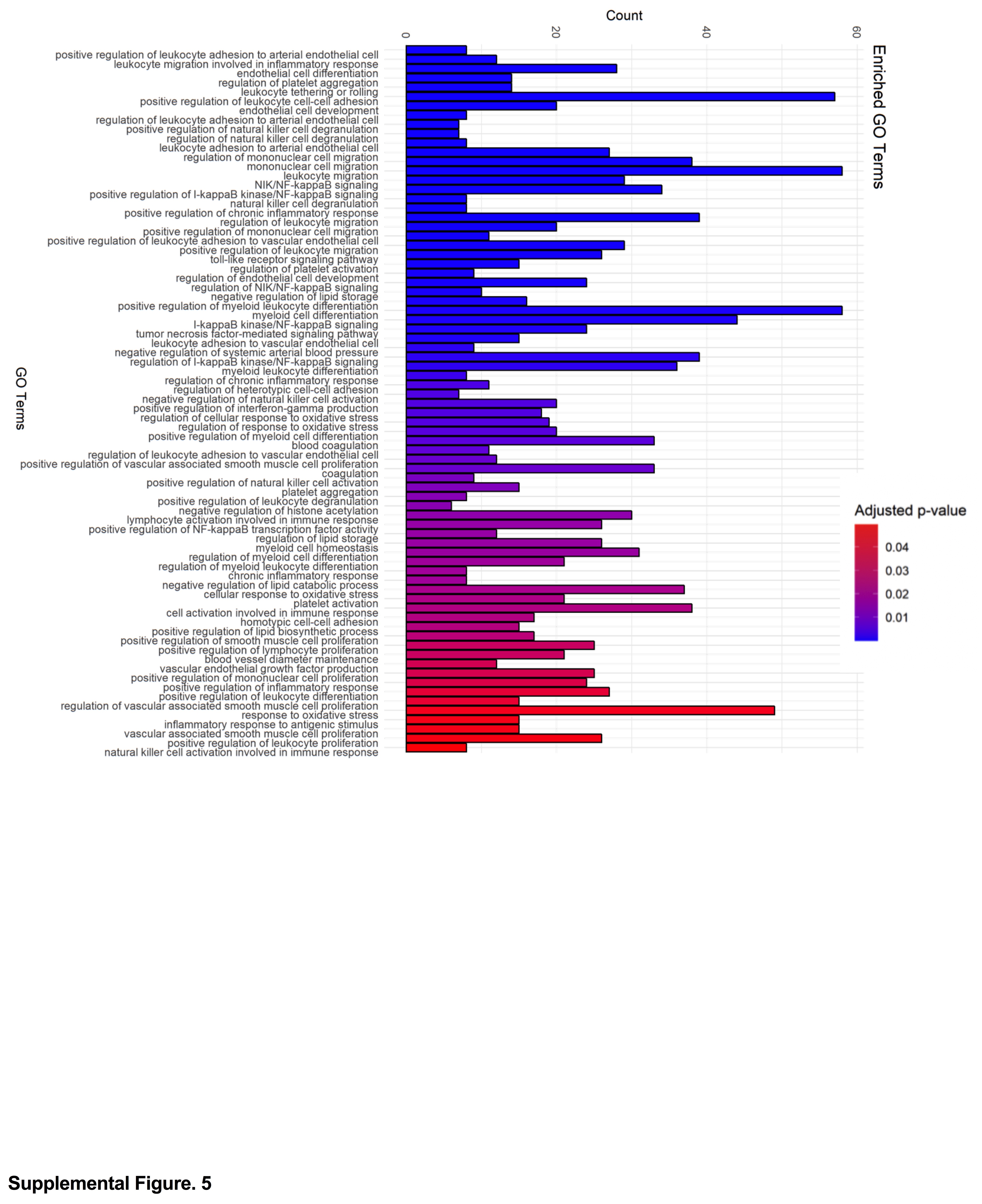

### Supplemental Figure 6

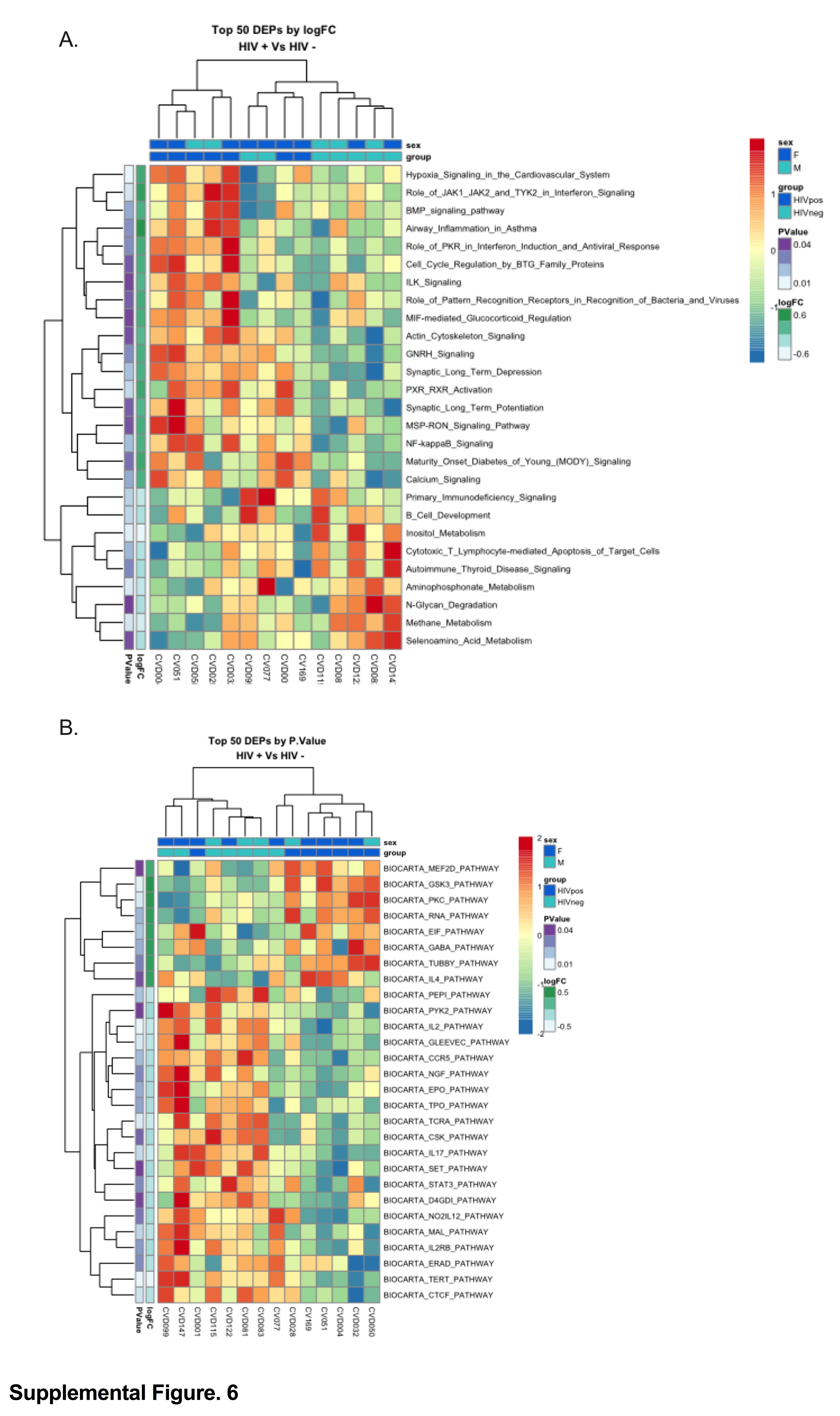
